# Antidepressant Treatment in Huntington’s Disease: Regional and Case-Control Variation

**DOI:** 10.1101/2025.08.01.25332817

**Authors:** Duncan Mclauchlan, Cheney Drew, Peter Holmans, Anne Rosser

## Abstract

**Objectives:**

1. Determine differences in frequency and choice of antidepressant for anxiety and depression between pwHD and controls.
2. Determine if regional variation affects antidepressant prescribing between pwHD and controls.

**Methods:** We used data from the observational cohort study ENROLL-HD. Clinical assessments and medication are recorded annually for 19540 pwHD and 6010 controls. We determined an episode of depression and anxiety as >1 for both severity and frequency Problem Behaviours Assessment(PBAs) depressed mood item and anxiety item respectively. We classed antidepressants as SSRI, SNRI, TCA, Unique, TeCA (tetracyclic antidepressant), NDRI (noradrenaline – dopamine reuptake inhibitor), phenylpiperazine. We used logistic models to determine the effect of case status on probability of antidepressant treatment and multinomial models to determine the effect of region and case status on antidepressant class.

**Results:** We found that rates of both depression(46.06% vs 32.42%) and anxiety(50.23% vs 37.47%) were higher in pwHD than controls. Even accounting for severity, pwHD were significantly more likely to receive an antidepressant than controls for both depression(OR 3.48,p<2×10^−16^) and anxiety(OR 4.34,p<2×10^−16^). Accounting for regional variation, pwHD were more likely than controls to receive a TeCA for depression(OR 2.1,p=1.2×10^−7^) or anxiety(OR 4.2,p=0.00025); and less likely to receive an NDRI for depression than controls(OR 0.78,p=0.022). There was substantial regional variation in antidepressant class selection for pwHD.

**Conclusions:** Anxiety and Depression are treated differently in pwHD and controls: the lack of an evidence base to justify this underscores the need for a clinical trial of antidepressants for depression and anxiety in pwHD.

## Background

Huntington’s disease is an autosomal dominant neurodegenerative disease, focussed on cortico-striatal networks. Depression and anxiety are very common in people with Huntington’s disease (pwHD). Large prevalence studies report lifetime rates of between 30 and 70% for depression and 30-60% for anxiety^1^, depending on the assessment tool and precise definition used, compared to rates in the wider population of between 10-20%^2^. It is likely that depression in HD is directly linked to HD neuropathology as previous work has shown that in cohorts of people at risk for HD blinded to their own genetic status, depression is higher in gene positive compared to gene negative groups^3,4^. In keeping with this, antidepressants are among the most frequently prescribed medication classes^5^ in ENROLL-HD^6^, a world-wide observational cohort study of pwHD and controls.

Previous work from our group has shown that the mechanisms underlying depression in HD differ from those leading to depression in control populations^7^. Furthermore, observational data suggests that the antidepressant classes with the best efficacy for depression in HD, also differed from those in the wider population^7,8^. To date there have been no randomised controlled trials (RCTs) of antidepressants for psychiatric symptoms in HD. Although several RCTs of antidepressants have reported depression as a secondary outcome, these excluded people with depression at study entry^9^. Case reports and open label studies^10,11^ report beneficial effects of antidepressants on depression in HD, but the rigour of these studies is very low.

Expert guidelines have been produced for treating a number of symptoms experienced by pwHD^12,13^. SSRIs are recommended as first line treatment of depression in HD. For depression in association with apathy (common in HD) then an activating antidepressant (such as fluoxetine or bupropion^14^) is recommended^13^. As sleep disorders (in particular insomnia) are common in HD, the recommendations are to prescribe a sedating antidepressant^12,13^ such as mirtazapine, phenylpiperazines or tricyclic antidepressants in the presence of significant sleep disturbance.

However, medication approvals differs across different regions^15^: for example in the UK bupropion is only approved by NICE for smoking cessation and not for depression. Further to this, medicine can be silo-ed by speciality and prescribing cultures can develop^16^. Antidepressant choice and threshold for prescription can also be influenced by deprivation, availability of non-drug interventions and social support^17^.However, this should not affect antidepressant choice between pwHD and controls in a given region. If there were regional differences in how pwHD and controls were treated (for example if pwHD in one region were more likely to be prescribed a SSRI than controls; but less likely to be prescribed an SSRI than controls in another) this would suggest a paucity in the evidence base for treating pwHD and underline the need for a clinical trial.

Using data from the ENROLL-HD study, which includes 25 550 participants in the most recent data release we performed the following analyses:

### Primary Analyses

1A. How frequently are antidepressants used to treat depression and anxiety in pwHD compared to controls?

1B. Accounting for regional variation, are depression and anxiety treated differently in participants with HD (pwHD) compared to controls; if so, does this follow established guidelines?

### Secondary Analyses

2. Is there regional variation in antidepressant class selection between pwHD and control participants which may indicate cultural prescribing patterns?

## Materials and Methods

We selected participants from the ENROLL-HD observational study. ENROLL-HD is a worldwide, observational study including people carrying the genetic mutation for Huntington’s disease, people at risk and family controls (confirmed gene negative or a family member not at risk of HD). The most recent data release (PDS6) includes 25 550 participants: 6008 family controls, and 19542 carrying the gene for HD. Participants genetic status is confirmed at baseline, and they then complete annual visits to assess motor features, function, cognition and psychiatric symptoms. Participants region (North America, Latin America, Europe, Australasia) is also recorded at baseline. Medication use is recorded annually: WHO ATC codes are used to categorise drug classes and indications for medication are recorded. We selected adult participants (age ≥18), either confirmed gene positive (at any stage of the disease: with or without symptoms), or confirmed gene negative/not at risk.

We determined if participants had experienced an episode of depression or anxiety based on the depressed mood and anxiety subscores from the Problem Behaviours Assessment (short form: PBAs^18^). The PBAs is a multidomain, clinician-scored neuropsychiatric instrument that scores severity (0-4) and frequency (0-4) of each individual symptom. We defined an episode of depression as having severity >1(at least “mild”) and frequency >1 (at least once weekly).

We included all antidepressant use (WHO ATC code N06A) in ENROLL-HD and categorised the indications for antidepressant use as follows: depression, anxiety, sleep, irritability/aggression, psychosis, other psychiatric symptom, motor symptom of HD, systemic symptom of HD, unclear. For example ‘melancholia’ was categorised as depression. To compare regional differences in antidepressant prescribing, we selected all antidepressants prescribed with an indication of depression, and as a secondary exploratory analysis antidepressants prescribed for anxiety.

We categorised antidepressant classes as previously described^7^: including separate categories for selective serotonin reuptake inhibitors (SSRIs), serotonin-noradrenaline reuptake inhibitors (SNRIs), noradrenaline-dopamine reuptake inhibitors (NDRIs), tricyclic antidepressants, phenylpiperazines, tetracyclic antidepressants, and agents with a unique mechanism of action (we included monamine oxidase inhibitors with unique agents, as their prevalence within ENROLL-HD was very small: <0.002%of all prescriptions). We included bupropion in a separate class (noradrenaline-dopamine reuptake inhibitor) as clinical practice would suggest this is an activating antidepressant (SSRIs can be activating or sedating), and separate classes for tetracyclic antidepressants (TeCAs) and phenylpiperazines as these classes both have sedating effects, and we wished to explore whether prescription patterns followed expert guidelines.

### Data Analysis

We fitted logistic models to determine the effect of case status (pwHD vs control) on the probability of receiving an antidepressant, including region, age, sex and symptom severity (mean PBA depression or anxiety product (severity*frequency) score^18^) as covariates. We used the multinom package in R^19^ (open source statistical software) to construct multinomial models in order to compare proportions of individuals prescribed the various classes of antidepressants across regions. Multinomial models compare relative frequencies across categories, using one category as a reference standard. To determine the effect of case status on antidepressant prescribing accounting for regional variation in prescribing patterns, we fitted a model in the whole population including antidepressant class as the dependent variable, and both region and case status (pwHD or Control) as independent variables, with co-variates age, sex and symptom severity. For our secondary analysis; we selected participants (pwHD and controls) from each region, and compared antidepressant choices between pwHD and controls, using an interaction between case status and region, with covariates of age, sex and symptom severity. The Bonferroni corrected alpha was 0.017, for the three analyses.

The funders of this work had no role in the concept, design, analysis or writing of this study. All study procedures in ENROLL-HD conformed to the declaration of Helskini, and all participants gave informed consent.

## Results

### Demographics

Table 1a&1b show the demographic breakdown for pwHD and controls receiving antidepressants for anxiety and depression. Although race and age were comparable across indications and disease status, there were higher numbers of females in the control group.

**Table 1a.** Demographics for Antidepressant Treated Cohort: Depression.

|  | Huntington's disease |  | Controls |  |
| --- | --- | --- | --- | --- |
| Age | 50.41 sd13.05 |  | 48.37 sd14.26 |  |
| Sex | 4720/8243 (57.26%) female |  | 664/871 (76.23%) female |  |
| Race | n | % | n | % |
| Black | 45 | 0.55% | 4 | 0.46% |
| Hispanic | 190 | 2.3% | 41 | 4.71% |
| Mixed | 66 | 0.8% | 7 | 0.8% |
| Native American | 19 | 0.23% | 8 | 0.92% |
| Other | 111 | 1.35% | 25 | 2.87% |
| White | 7770 | 94.26% | 780 | 89.55% |
| Not Recorded | 42 | 0.51% | 6 | 0.69% |

**Table 1b:** Demographics for Antidepressant Treated Cohort: Anxiety.

|  | Huntington's disease |  | Controls |  |
| --- | --- | --- | --- | --- |
| Age | 48.68 sd13.46 |  | 47.1 sd14.75 |  |
| Sex | 1128/1994 (56.56%) female |  | 242/305 (79.34%) female |  |
| Race | n | % | n | % |
| Black | 10 | 0.5% | 1 | 0.35% |
| Hispanic | 52 | 2.61% | 13 | 4.26% |
| Mixed | 16 | 0.8% | 2 | 0.66% |
| Native American | 4 | 0.2% | 3 | 0.98% |
| Other | 40 | 2% | 5 | 1.64% |
| White | 1865 | 93.53% | 281 | 92.13% |
| Not Recorded | 7 | 0.35% |  |  |

### Differences in Frequency of Antidepressant Treatment between HD and Controls

A total of 871/6008 control participants (Europe: 328, Australasia: 42, Latin America: 27, North America: 474) and 8243/19542 pwHD (Europe: 5366, Australasia: 286, Latin America: 135, North America: 2456) received an antidepressant for depression. Whilst 305/6008 control participants (Europe: 92, Australasia: 8, Latin America: 4, North America: 201) and 1994/19542 pwHD (Europe: 1061, Australasia: 79, Latin America: 14, North America: 840) received an antidepressant for anxiety.

In keeping with previous study findings, lifetime prevalence of anxiety (one episode of PBAs anxiety severity and frequency score both >1)(50.23% pwHD; 37.47% controls: χ-squared = 301.35, df = 2, p-value < 2×10^−16^) and depression (one episode of PBAs depressed mood severity and frequency score both >1)(9001/19540 (46.06%) pwHD; 1949/6003 (32.42%) controls: χ-squared = 350.26, df = 2, p-value < 2×10^−16^) was higher in pwHD than controls.

For both anxiety and depression, rates of antidepressant treatment were higher in pwHD than controls. Of the 9001 pwHD who experienced an episode of depression during the study, 58.35% (5252) received an antidepressant; whilst of the 1387 controls with an episode of depression, 562 (28.84%) received an antidepressant for depression. Similarly, of the pwHD experiencing anxiety (9815/19540; 50.23%); 1418 (14.44%) of them received an antidepressant; whilst of the 2252 controls experiencing anxiety, 222 (9.86%) were treated with an antidepressant. Fitting logistic models (supplementary data Tables S1A&S1B), to determine the effect of case status on antidepressant treatment with accounting for age, sex region and symptom severity (average PBAs symptom score at each visit) showed that pwHD were significantly more likely to receive an antidepressant than controls for both depression (pseudo R^2^=0.051; odds ratio 3.48, (confidence intervals(CIs) 3.12,3.88),p<2×10^−16^) and anxiety (pseudo R^2^=0.068; odds ratio 4.34 CIs(3.9,4.82),p <2×10^−16^). The logistic models including an interaction between case status and region (supplementary data Tables S1C&S1D) did not show any significant interactions: there is no region where the probability of pwHD receiving antidepressant treatment relative to that of controls is substantially different from the others.

Differences in Antidepressant Choice for Depression and Anxiety Between HD and Controls (Supplementary Data Tables S2A&B; Figure 1a&b)

**Figure 1.**
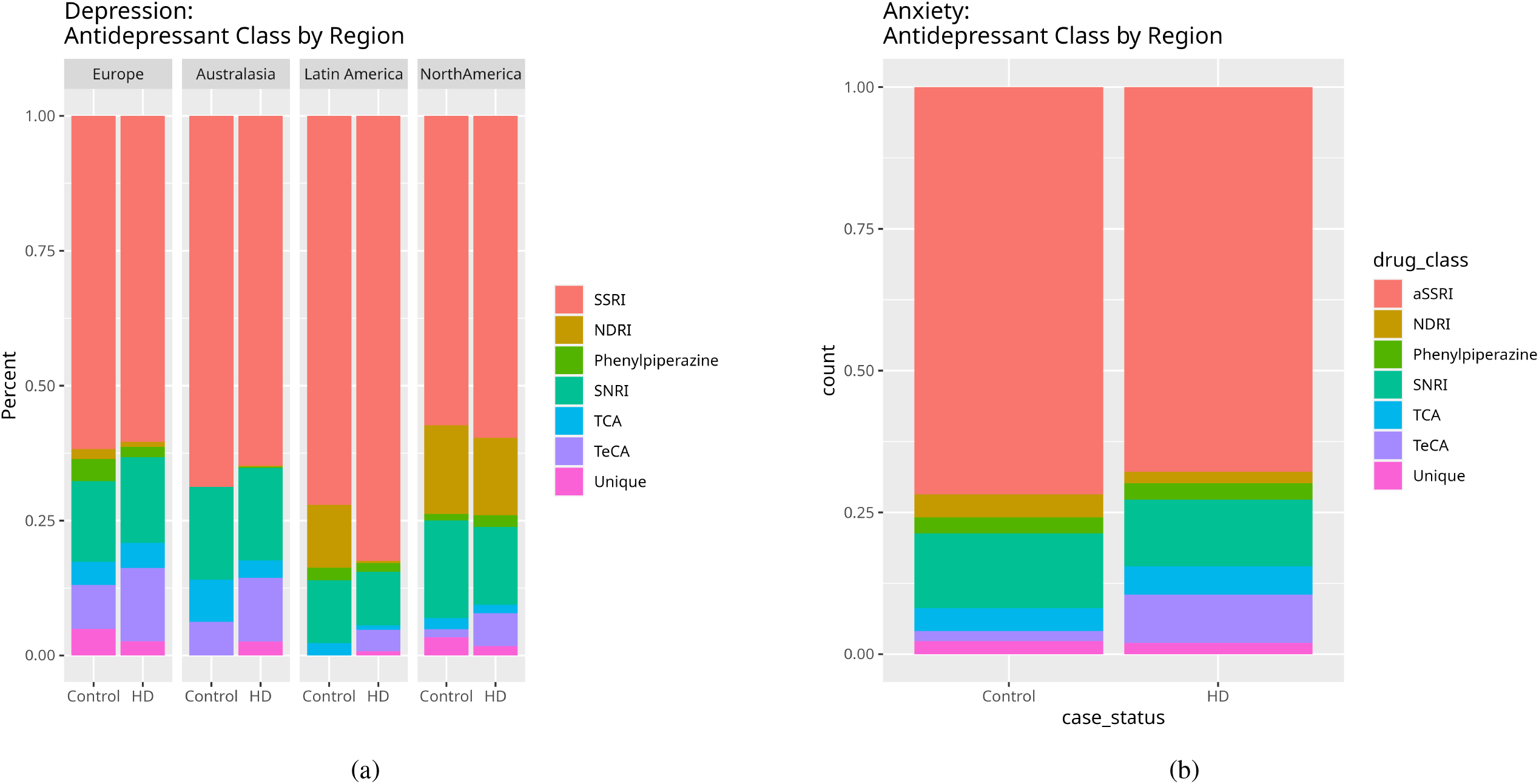
Antidepressant Choice for a) Depression and b) Anxiety.

Antidepressant guidelines for depression and anxiety in HD recommend considering activating antidepressants in the presence of co-morbid apathy (such as NDRIs) and sedating antidepressants with co-morbid insomnia (TeCAs, Phenylpiperazines, TCAs). Rates of SSRI prescription were similar in pwHD and controls for depression (Table 2a: 60.67% and 60.11% respectively). Relative to SSRIs, depression in pwHD was less likely to be treated with a unique agent (Table 2a: pwHD 2.36%, controls 3.7%; odds ratio 0.56 SE(0.094) p= 3.08 x10^−4^) or NDRI (Table 2a: pwHD 4.2%, controls 9.83%; odds ratio 0.77 SE(0.084), p= 0.015) compared to controls; whilst TeCAs were more frequently prescribed (Table 2a: pwHD 11.59%, controls 4.25%; odds ratio 2.1 SE(0.3), p=5.4×10^−7^) for depression in pwHD than in controls. Rates of SSRI prescription were slightly lower in pwHD than controls for depression (Table 2a: 67.82% and 71.83% respectively).In keeping with the findings for depression treatment, relative to SSRIs, pwHD were more likely to receive a TeCA than controls (Table 2a: pwHD 8.49%, controls 1.78%; odds ratio 4.2 SE(1.6), p=0.00043) for anxiety.

**Table 2a.** Antidepressant Choice for Depression by Region:

| HUNTINGTON'S DISEASE |  |  |  |  |  |
| --- | --- | --- | --- | --- | --- |
| Drug Class | Europe | Australasia | Latin America | North America | Total |
|  | percent (N) | percent (N) | percent (N) | percent (N) | percent (N) |
| SSRI | 60.38 (6947) | 64.82 (328) | 82.47 (207) | 59.64 (2367) | 60.67 (9849) |
| NDRI | 0.94 (108) | 0.20 (1) | 0.40 (1) | 14.39 (571) | 4.2 (681) |
| Phenylpiperazine | 1.92 (221) | 0.20 (1) | 1.59 (4) | 2.12 (84) | 1.91 (310) |
| SNRI | 15.86 (1825) | 17.19 (87) | 9.96 (25) | 14.44 (573) | 15.46 (2510) |
| TCA | 4.66 (536) | 3.16 (16) | 0.80 (2) | 1.59 (64) | 3.81 (618) |
| TeCA | 13.63 (1568) | 11.86 (60) | 3.98 (10) | 6.12 (243) | 11.59 (1881) |
| Unique | 2.61 (300) | 2.57 (13) | 0.80 (2) | 1.71 (68) | 2.36 (383) |
| CONTROL |  |  |  |  |  |
| Drug Class | Europe | Australasia | Latin America | North America | Total |
|  | percent (N) | percent (N) | percent (N) | percent (N) | percent (N) |
| SSRI | 61.76 (302) | 68.75 (44) | 72.09 (31) | 57.33 (387) | 60.11 (764) |
| NDRI | 1.84 (9) | 0.00 (0) | 11.63 (5) | 16.44 (111) | 9.83 (125) |
| Phenylpiperazine | 4.09 (20) | 0.00 (0) | 2.33 (1) | 1.19 (8) | 2.28 (29) |
| SNRI | 14.93 (73) | 17.19 (11) | 11.63 (5) | 18.07 (122) | 16.6 (211) |
| TCA | 4.29 (21) | 7.81 (5) | 2.33 (1) | 2.07 (14) | 3.23 (41) |
| TeCA | 8.18 (40) | 6.25 (4) | 0.00 (0) | 1.48 (10) | 4.25 (54) |
| Unique | 4.91 (24) | 0.00 (0) | 0.00 (0) | 3.41 (23) | 3.7 (47) |

**Table 2b:** Antidepressant Choice for Anxiety by Region:

| HUNTINGTON'S DISEASE |  |  |  |  |  |
| --- | --- | --- | --- | --- | --- |
| Drug Class | Europe | Australasia | Latin America | North America | Total |
|  | percent (N) | percent (N) | percent (N) | percent (N) | percent (N) |
| SSRI | 65.52 (1142) | 70.15 (94) | 89.29 (25) | 70.41 (847) | 67.82 (2108) |
| NDRI | 0.29 (5) | 0.00 (0) | 0.00 (0) | 4.82 (58) | 2.03 (63) |
| Phenylpiperazine | 2.81 (49) | 0.00 (0) | 0.00 (0) | 3.41 (41) | 2.9 (90) |
| SNRI | 11.70 (204) | 8.21 (11) | 3.57 (1) | 12.47 (150) | 11.78 (366) |
| TCA | 6.94 (121) | 2.24 (3) | 7.14 (2) | 2.41 (29) | 4.99 (155) |
| TeCA | 10.27 (179) | 14.93 (20) | 0.00 (0) | 5.40 (65) | 8.49 (264) |
| Unique | 2.47 (43) | 4.48 (6) | 0.00 (0) | 1.08 (13) | 1.99 (62) |
| CONTROL |  |  |  |  |  |
| Drug Class | Europe | Australasia | Latin America | North America | Total |
|  | percent (N) | percent (N) | percent (N) | percent (N) | percent (N) |
| SSRI | 71.77 (89) | 88.89 (8) | 83.33 (5) | 70.98 (181) | 71.83 (283) |
| NDRI | 0.00 (0) | 0.00 (0) | 0.00 (0) | 6.27 (16) | 4.1 (16) |
| Phenylpiperazine | 1.61 (2) | 0.00 (0) | 16.67 (1) | 3.14 (8) | 2.79 (11) |
| SNRI | 13.71 (17) | 0.00 (0) | 0.00 (0) | 13.73 (35) | 13.2 (52) |
| TCA | 7.26 (9) | 11.11 (1) | 0.00 (0) | 2.35 (6) | 4.1 (16) |
| TeCA | 3.23 (4) | 0.00 (0) | 0.00 (0) | 1.18 (3) | 1.78 (7) |
| Unique | 2.42 (3) | 0.00 (0) | 0.00 (0) | 2.35 (6) | 2.28 (9) |

Regional Differences in Antidepressant Choice for Depression Between HD and Controls (Supplementary Data Table S3(ii))

There was very little consistency across regions in whether depression should be treated differently in pwHD compared to controls, and if so in what way. The multinomial model, with an outcome of antidepressant class and an interaction between region and case status (to determine if differences between antidepressant class selection between cases and controls varied across regions) showed a number of significant findings for depression treatment. Use of phenylpiperazines rather than SSRIs in pwHD relative to controls is significantly higher in North American than in Europe (Europe pwHD 1.29% Europe controls 4.09%; North America pwHD 2.12% North America Controls 1.19%; OR 3.3 SE(0.0056),p=0.0071) and the same disparity was noted for TeCAs (Europe pwHD 13.63% Europe controls 8.18%; North America pwHD 11.59%, North America Controls 4.25%; OR 2.3 SE(<2×10^−16^),p=0.021). The N for some antidepressant classes in controls was very small in Latin America and Australasia (Table 2a). Nonetheless, antidepressant class choice between cases and controls showed marked differences to Europe, for all antidepressant classes except for SNRIs (Supplementary Data Table S3(ii)).

## Discussion

In this work, we report a number of significant findings. Firstly, depression and anxiety were treated differently in pwHD and controls: pwHD were more likely to receive an antidepressant than controls, and were more likely to receive a TeCA (for depression and anxiety) and less likely to receive a NDRI (for depression). This finding suggests that although sedating antidepressants are being appropriately used more often in pwHD, activating antidepressants are not. In keeping with this, there is substantial regional variation in treatment choices for depression between pwHD and controls.

We found that antidepressants were much more frequently prescribed to pwHD than controls, even after correcting for symptom severity. The reason underlying this disparity is unclear. It may reflect the higher rates of suicide in pwHD: with depressive symptoms being treated more aggressively, however the disparity was also seen in treatment of anxiety. There may be a dearth of psychological services for pwHD compared to controls^20^, leading to higher frequency of medical treatment. Alternatively, treating clinicians may feel antidepressants are more effective in pwHD than in controls. The lack of an evidence base to justify the disparity in treatment underscores the need for a clinical trial.

Multiple opinion-led clinical guidelines for treating depression in HD^12,13^ recommend consideration of activating and sedating antidepressants in the presence of relevant co-morbidities (apathy, and insomnia respectively). Consequently we expected to find higher rates of both types of antidepressant. However, the data suggests these guidelines are not being adhered to in clinical practice. This disparity cannot be explained by different regulatory regimes, as the regions where NDRIs are most commonly prescribed (Latin America and North America), had lower rates of NDRI use for depression in pwHD compared to controls. This may reflect concerns over seizure risk (a known side effect of bupropion), as seizures can be more common, particularly in juvenile onset HD^21^. Alternatively owing to the behavioural changes in HD there may be concern amongst treating clinicians about agitation or impulsive behaviour which can be exacerbated by activating antidepressants^14^. The preference for TeCAs over TCAs or phenylpiperazines may relate to the cholinergic side effects seen with the latter two classes.

The data also show significant disparities across regions regarding best practice in treating depression in pwHD compared to controls. For example NDRIs were more frequently used for depression in pwHD compared to controls in Australasia and less frequently used in Latin America. It is not clear what underlies these differences, but it is likely to reflect the lack of an evidence base regarding antidepressant efficacy for anxiety and depression in HD, underscoring the pressing need for a head to head randomised clinical trial of antidepressants for depression in HD.

This work does have some limitations: ENROLL-HD does not release individual country or site data so it is difficult to make more fine-grained analyses on regulatory regimes across nations and individual practices. The indication recorded, is based on participant report and may not always be correct (participants may lack insight into irritability, and may believe the antidepressant is for anxiety for example). However, the large sample size may mitigate this to some extent. Although there was a higher proportion of women in the control group, we corrected for sex in the analyses, and evidence from large registries suggests that women are more likely to receive an antidepressant than men^22^. We were not able to make any inferences regarding irritability, as the number of control participants prescribed antidepressants for this indication was negligible.

In this work, we have demonstrated that pwHD who experience depression are being treated differently from controls, but not in keeping with international guidelines. We show that the disparity in antidepressant choice for pwHD compared to controls varies across regions. These disparities underscore the pressing need for an efficacy trial of antidepressants for depression in HD.

## Data Availability

All data from Enroll-HD are available on application for any qualified researcher through the scientific review committee of the CHDI foundation at https://enroll-hd.org/

## Funding

ENROLL-HD and TRACK-HD were funded by CHDI Foundation. Dr McLauchlan is funded by a Health and Care Research Wales Research Time Award(NHS.RTA-22-15). Cheney Drew is funded by Health Care Research Wales, Michael J Fox Foundation and Jacques and Gloria Gossweiler Foundation. All other authors are funded by Cardiff University. The funders played no role in the design, conduct or analysis of this work.

**Table S1A:**
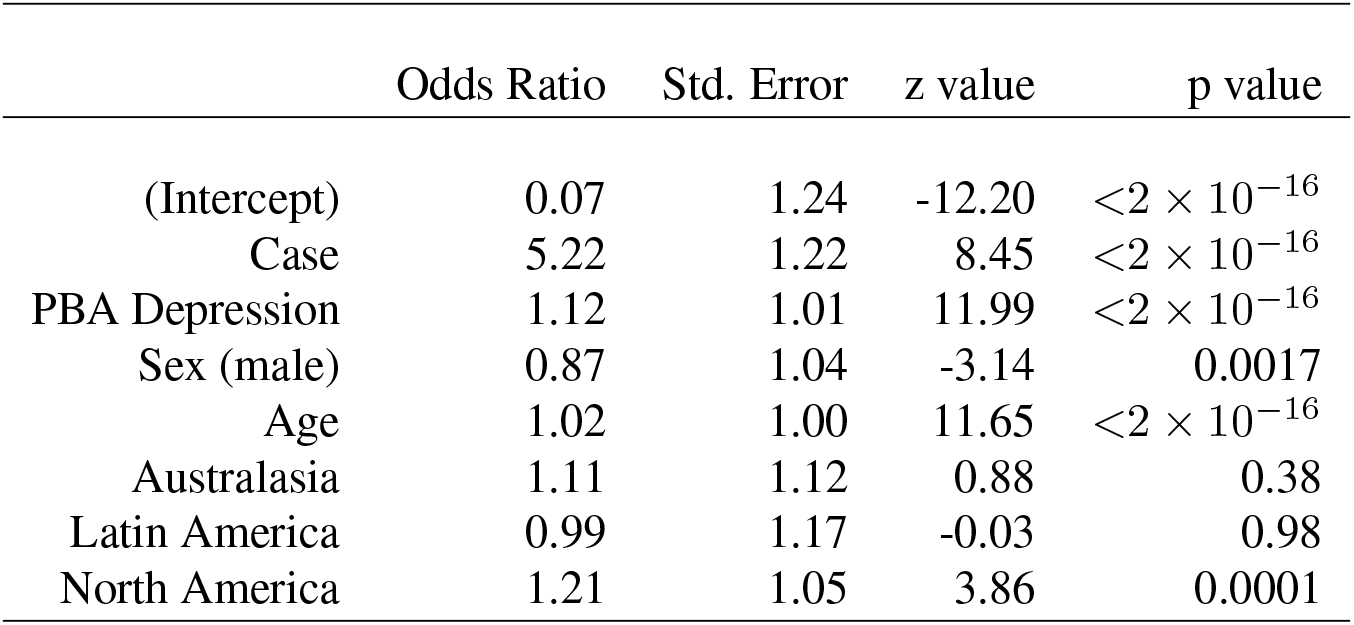
Differences in Antidepressant Treatment Probability for Depression between HD and Controls.

**Table S1B:** Differences in Antidepressant Treatment Probability for Depression between HD and Controls by Region.

|  | Odds Ratio | Std. Error | z value | p value |
| --- | --- | --- | --- | --- |
| (Intercept) | 0.06 | 1.45 | -7.49 | $6.73 \times 10^{-14}$ |
| Case | 6.01 | 1.44 | 4.95 | $7.59 \times 10^{-7}$ |
| Australasia | 0.93 | 3.22 | -0.06 | 0.95 |
| Latin America | 2.84 | 4.37 | 0.71 | 0.48 |
| North America | 1.47 | 1.54 | 0.89 | 0.37 |
| PBA Depression | 1.12 | 1.01 | 11.98 | $<2 \times 10^{-16}$ |
| Sex (male) | 0.87 | 1.04 | -3.14 | 0.0017 |
| Age | 1.02 | 1.00 | 11.65 | $<2 \times 10^{-16}$ |
| Case: Australasia | 1.20 | 3.24 | 0.15 | 0.88 |
| Case: Latin America | 0.35 | 4.41 | -0.71 | 0.48 |
| Case: North America | 0.82 | 1.55 | -0.45 | 0.65 |

**Table S1C:** Differences in Antidepressant Treatment Probability for Anxiety between HD and Controls.

|  | Odds Ratio | Std. Error | z value | p value |
| --- | --- | --- | --- | --- |
| (Intercept) | 0.07 | 1.22 | -13.57 | $<2 \times 10^{-16}$ |
| Case | 6.79 | 1.19 | 10.87 | $<2 \times 10^{-16}$ |
| PBA Anxiety | 1.06 | 1.01 | 7.49 | $<2 \times 10^{-16}$ |
| Sex (male) | 0.87 | 1.04 | -3.07 | 0.0021 |
| Age | 1.02 | 1.00 | 11.39 | $<2 \times 10^{-16}$ |
| Australasia | 1.05 | 1.12 | 0.42 | 0.67 |
| Latin America | 1.03 | 1.17 | 0.22 | 0.83 |
| North America | 1.19 | 1.05 | 3.48 | 0.0005 |

**Table S1D:** Differences in Antidepressant Treatment Probability for Anxiety between HD and Controls by Region.

|  | Odds Ratio | Std. Error | z value | p value |
| --- | --- | --- | --- | --- |
| (Intercept) | 0.07 | 1.38 | -8.47 | $<2 \times 10^{-16}$ |
| Case | 6.98 | 1.36 | 6.27 | $3.58 \times 10^{-10}$ |
| Australasia | 1.85 | 2.42 | 0.70 | 0.49 |
| Latin America | 1.67 | 3.39 | 0.42 | 0.67 |
| North America | 1.18 | 1.46 | 0.44 | 0.66 |
| PBA Anxiety | 1.06 | 1.01 | 7.49 | $6.89 \times 10^{-14}$ |
| Sex (male) | 0.87 | 1.04 | -3.07 | 0.0021 |
| Age | 1.02 | 1.00 | 11.39 | $<2 \times 10^{-16}$ |
| Case: Australasia | 0.56 | 2.44 | -0.65 | 0.52 |
| Case: Latin America | 0.61 | 3.43 | -0.40 | 0.69 |
| Case: North America | 1.01 | 1.46 | 0.02 | 0.99 |

**Table S2A:**
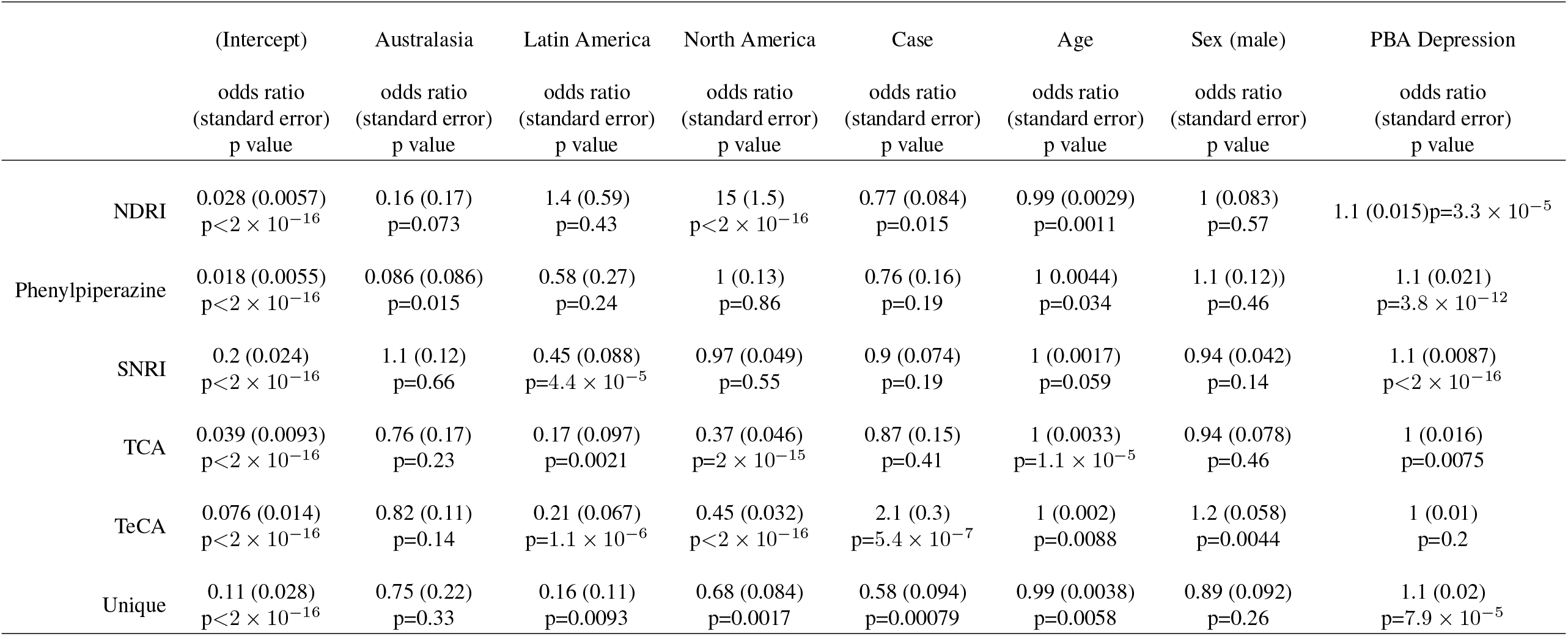
Differences in Antidepressant Class Prescribig for Depression.

**Table S2B:**
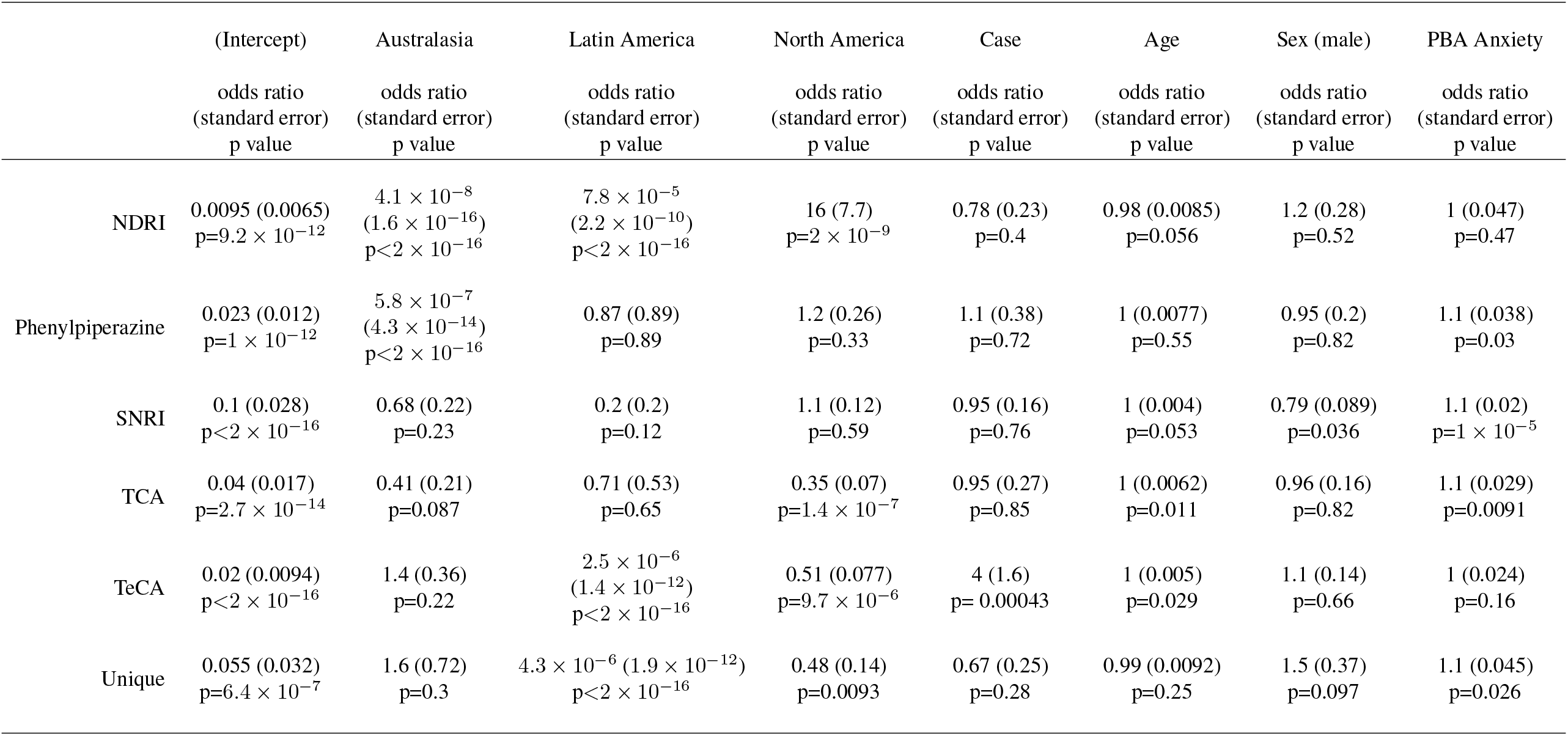
Differences in Antidepressant Class Prescribing for Anxiety.

**Table S3(i):** Differences in Antidepressant Class Prescribing for Depression Effect of Region on Antidepressant Choice for Depression.

|  | (Intercept) | Australasia | Latin America | North America | Case | Age | Sex (male) | PBA Depression |
| --- | --- | --- | --- | --- | --- | --- | --- | --- |
|  | odds ratio<br>(standard error)<br>p value | odds ratio<br>(standard error)<br>p value | odds ratio<br>(standard error)<br>p value | odds ratio<br>(standard error)<br>p value | odds ratio<br>(standard error)<br>p value | odds ratio<br>(standard error)<br>p value | odds ratio<br>(standard error)<br>p value | odds ratio<br>(standard error)<br>p value |
| NDRI | 0.039 (0.0079)<br>$p < 2 \times 10^{-16}$ | $6.7 \times 10^{-5}$<br>($6.7 \times 10^{-5}$)<br>$p < 2 \times 10^{-16}$ | 5.1 (2.2)<br>$p = 0.0057$ | 10 (1.1)$p = 6.7 \times 10^{-11}$ | 0.54 (0.06)<br>$p = 0.082$ | 0.99 (0.0029)<br>$p = 0.0011$ | 1 (0.083)<br>$p = 0.56$ | 1.1 (0.015)<br>$p = 4.3 \times 10^{-5}$ |
| Phenylpiperazine | 0.027 (0.0035)<br>$p < 2 \times 10^{-16}$ | $1.2 \times 10^{-6}$<br>($2.4 \times 10^{-7}$)<br>$p < 2 \times 10^{-16}$ | 0.44 (0.0019)<br>$p = 0.43$ | 0.35 (0.039)<br>$p = 0.013$ | 0.48 (0.0087)<br>$p = 0.0022$ | 1 (0.12)<br>$p = 0.03$ | 1.1 (0.13)<br>$p = 0.48$ | 1.1 (0.22)<br>$p = 9 \times 10^{-12}$ |
| SNRI | 0.16 (0.0071)<br>$p < 2 \times 10^{-16}$ | 1.1 (0.0084)<br>$p = 0.88$ | 0.61(0.15)<br>$p = 0.32$ | 1.4 (0.32)<br>$p = 0.045$ | 1.1 (0.65)<br>$p = 0.43$ | 1 (0.13)<br>$p = 0.067$ | 0.94 (0.16)<br>$p = 0.14$ | 1.1 (0.0035)<br>$p < 2 \times 10^{-16}$ |
| TCA | 0.031 (0.0044)<br>$p < 2 \times 10^{-16}$ | 1.7 (0.55)<br>$p = 0.32$ | 0.45 (0.032)<br>$p = 0.44$ | 0.53 (0.077)<br>$p = 0.072$ | 1.1 (0.0022)<br>$p = 0.73$ | 1 (0.051)<br>$p = 1.3 \times 10^{-5}$ | 0.94 (0.0096)<br>$p = 0.47$ | 1 (0.26)<br>$p = 0.0074$ |
| TeCA | 0.095 (0.016)<br>$p < 2 \times 10^{-16}$ | 0.7 (0.0027)<br>$p = 0.51$ | $5.6 \times 10^{-6}$<br>($5.8 \times 10^{-7}$)<br>$p < 2 \times 10^{-16}$ | 0.2 (0.0036)<br>$p = 7.6 \times 10^{-6}$ | 1.6<br>($< 2 \times 10^{-16}$)<br>$p = 0.0038$ | 1<br>($< 2 \times 10^{-16}$)<br>$p = 0.0079$ | 1.2<br>($< 2 \times 10^{-16}$)<br>$p = 0.0046$ | 1<br>($< 2 \times 10^{-16}$)<br>$p = 0.21$ |
| Unique | 0.11<br>($< 2 \times 10^{-16}$)<br>$p = 7.5 \times 10^{-15}$ | $1.1 \times 10^{-5}$<br>($< 2 \times 10^{-16}$)<br>$p < 2 \times 10^{-16}$ | $6.6 \times 10^{-6}$<br>($< 2 \times 10^{-16}$)<br>$p < 2 \times 10^{-16}$ | 0.79<br>($< 2 \times 10^{-16}$)<br>$p = 0.44$ | 0.58<br>($< 2 \times 10^{-16}$)<br>$p = 0.014$ | 0.99<br>($< 2 \times 10^{-16}$)<br>$p = 0.0055$ | 0.89<br>($< 2 \times 10^{-16}$)<br>$p = 0.26$ | 1.1<br>($< 2 \times 10^{-16}$)<br>$p = 6.2 \times 10^{-5}$ |

**Table S3(ii):** Differences in Antidepressant Class Prescribing for Depression Effect of Region on Antidepressant Choice for Depression.

| Interactions |  |  |  |  |
| --- | --- | --- | --- | --- |
|  | Australasia | Latin America | North America |  |
|  | odds ratio<br>(standard error)<br>p value | odds ratio<br>(standard error)<br>p value | odds ratio<br>(standard error)<br>p value |  |
| NDRI | $3 \times 10^3$<br>($9.2 \times 10^2$)<br>$p < 2 \times 10^{-16}$ | 0.056 (0.056)<br>p=0.014 | 1.5 (0.69)<br>p=0.27 | |
| Phenylpiperazine | $8.9 \times 10^4$<br>($4.5 \times 10^3$)<br>$p < 2 \times 10^{-16}$ | 1.3 (0.11)<br>p=0.81 | 3.3 (0.0056)<br>p=0.0071 | |
| SNRI | 1 (0.084)<br>p=0.97 | 0.72 (0.011)<br>p=0.54 | 0.67 (0.12)<br>p=0.021 |  |
| TCA | 0.39 (0.11)<br>p=0.11 | 0.29 (0.2)<br>p=0.32 | 0.67 (0.083)<br>p=0.29 |  |
| TeCA | 1.2<br>($< 2 \times 10^{-16}$)<br>p=0.77 | $3.9 \times 10^4$<br>($< 2 \times 10^{-16}$)<br>$p < 2 \times 10^{-16}$ | 2.3<br>($< 2 \times 10^{-16}$)<br>p=0.021 | |
| Unique | $8.6 \times 10^4$<br>($< 2 \times 10^{-16}$)<br>$p < 2 \times 10^{-16}$ | $3.1 \times 10^4$<br>($< 2 \times 10^{-16}$)<br>$p < 2 \times 10^{-16}$ | 0.82<br>($< 2 \times 10^{-16}$)<br>p=0.55 | |

